# An integrated rural health system baseline assessment of COVID-19 preparedness in Siaya Kenya

**DOI:** 10.1101/2021.02.07.21251312

**Authors:** Neema Kaseje, Dan Kaseje, Kennedy Oruenjo, Penina Ocholla Odhiambo, Margaret Kaseje, Stephen Achola, Marcel Tanner, Andrew Haines

## Abstract

**Objective:** Our aim was to assess Siaya county COVID-19 preparedness at community and health facility levels and measure baseline household prevalences of fever and cough.

**Design:** There was retrospective and prospective data collection using standard tools. We determined the prevalence of fever and cough in households. We evaluated household knowledge about COVID-19 prevention and adherence to preventive measures. We evaluated the presence of a workforce, essential infrastructure and equipment needed for COVID-19 case management, and the availability of essential maternal and child health services in health facilities.

**Setting:** Siaya in rural Western Kenya

**Participants:** households and health facilities in Siaya

**Results:** We visited 19’474 households and assessed 152 facilities. The prevalences of fever and cough ranged from 1.4% to 4.3% and 0.2 to 0.8% respectively; 97% and 98% of households had not received a guest from nor travelled outside Siaya respectively; 97% knew about frequent handwashing, 66% knew about keeping distance, and 80% knew about wearing a mask; 63% washed their hands countless times; 53% remained home; and 74% used a mask when out in public. The health facility assessment showed: 93.6% were dispensaries and health centers; 90.4% had nurses; 40.5% had oxygen capacity; 13.5% had pulse oximeters; and 2 ventilators were available; 94.2% of facilities did not have COVID-19 testing kits; 94% and 91% of facilities continued to provide antenatal care and immunization services respectively. Health care worker training in COVID-19 had been planned.

**Conclusions:** Household prevalence of fever and cough was low suggesting Siaya had not entered the active community transmission phase in June 2020. Our assessment revealed a need for training in COVID-19 case management, and a need for basic equipment and supplies including pulse oximeters and oxygen. Future interventions should address these gaps.

**Strengths and limitations:**

- This study provides an example of how to successfully carry out an integrated rural health system baseline assessment of COVID-19 preparedness; an approach that would be useful for any country experiencing COVID-19 with a significant rural population.
- Some of our data were retrospective in nature and therefore vulnerable to multiple sources of bias including: recall bias and misclassification.

**Clinical Trial registration:** Clinicaltrials.gov NCT04501458 5/8/2020

**Protocol:** The full protocol has been accepted for publication: Kaseje N, Kaseje D, Oruenjo K, Milambo J and Kaseje M: Engaging community health workers, technology, and youth in the COVID-19 response with concurrent critical care capacity building: A protocol for an integrated community and health system intervention to reduce mortality related to COVID-19 infection in Western Kenya. Wellcome Open Research.

**Ethical review approvals:** received from the University of Nairobi Ethics Review Committee and Jaramogi Oginga Odinga Teaching and Referral Hospital Ethics Review Committee (**approval number IERC/JOOTR/219/20**)

## Introduction

During the initial phase of the pandemic in May and June 2020, globally close to 11 million COVID-19 cases had been declared with more than 500’000 deaths (1). There were growing numbers in sub Saharan Africa: Kenya, had reported more than 6000 cases with 149 deaths (1). The highest numbers of cases were in Nairobi and Mombasa and movements in and out of these regions were restricted (2). Although sub-saharan Africa has experienced pandemics in the past including the HIV-AIDs pandemic and highly infectious outbreaks such as Ebola, little is known about its rural health systems’ capacity to prepare for and respond to the COVID-19 pandemic (3).

Fifty nine percent of the population in Africa is rural (4). In Kenya, 72% of the population is rural (4). Rural populations face greater challenges when accessing health services because of an inadequate workforce density, inadequate number of facilities, and inadequate infrastructure (including roads and communications) (5). These challenges faced by rural populations are likely to persist during the COVID-19 pandemic and an approach that is tailored to the rural context is necessary.

As early as 2007, a call was made to strengthen preparedness for a highly pathogenic influenza like pandemic in Africa (6). More recently, the COVID-19 pandemic has led to the realization for the need to strengthen primary health care systems (7). Community health workers (CHWs) play a critical role in primary health care systems in sub Saharan Africa, and their role in pandemics is increasingly being recognized: Corburn et al recommended immediate training and deployment of CHWs to address COVID-19 in informal settlements of the global south (8). In addition, Ballard M et al highlight the need to prioritize CHWs in the COVID-19 response emphasizing: protection of healthcare workers, interruption of the virus, maintenance of existing health services while surging their capacity, and shielding the most vulnerable from socio-economic shocks (9).

Not only do CHWs play a critical role in educating and promoting good health among household members, but there is empirical evidence demonstrating the validity and reliability of CHW collected data for planning and policy in Western Kenya (10,11). This is important in the Siaya context, given that community-based health care has been in place since the establishment of the Saradidi rural health project in Rarieda (12,13).

Given that an ongoing COVID-19 pandemic is likely to put a strain on the limited rural health workforce, health facilities, and basic infrastructure and equipment available, a rapid assessment of preparedness is necessary for planning and formulating an effective COVID-19 intervention.

Thus, our first goal was to rapidly assess community based preparedness and health behavior related to COVID-19 prevention using CHWs accompanied by youth and digital tools. Our second goal was to examine the health system’s capacity to manage suspected, and confirmed cases of COVID-19, and its capacity to maintain essential health services during the COVID-19 response.

## Methods

### Setting

Siaya is located in Western Kenya and is bordered by Busia that borders Uganda, Kakamega, Vihiga and Kisumu. It has a total surface area of 2500 km squared with a population of 993’000 in the six sub-counties. Seventy five percent of the population is rural; 27% live on less than $1 per day. Most are subsistence farmers. Siaya has one of the worst health indicators in the country: infant mortality rate is 111 per 1000 live births compared to 49 nationally; maternal mortality rate is 695 per 100’000 compared to a national rate of 488 per 100000. The county has a total of 175 health facilities with less than 10 intensive care unit beds.

We started baseline activities in Rarieda subcounty and later proceeded to 4 additional subcounties. The sixth sub-county was not ready for baseline activities and was to be included later. At the household level, we assessed knowledge of COVID-19 prevention, health behavior related to COVID19 prevention, prevalence of fever and other COVID-19 symptoms, and utilization of essential maternal and child health services. In addition, we assessed community health unit (CHU) functionality. A CHU serves approximately 1,000 households (or 5,000 people).

At the health system level, we assessed the availability and the composition of the health workforce, the availability of key equipment including pulse oximeters; the availability of key supplies including oxygen; and the presence of operating room theaters that could be reorganized to build critical care capacity. Furthermore, we assessed the presence of personal protective equipment; the presence of training in basic critical care and the management of severe respiratory infection including COVID19; and the availability of essential maternal and child health services.

The full protocol has been previously described (14).

### Community level

First, we conducted training sessions to introduce the digital household survey to community health workers, youth, and community health assistants (CHAs). During the training session, and after introducing COVID-19 as a disease process and its potential impact on communities, each item in the digital tools was reviewed by training facilitators. Participants were given the chance to ask clarifying questions. Furthermore, participants were given the opportunity to practice using the digital tool. During these sessions, infrared contactless thermometers were distributed. Facilitators examined and made sure the thermometers were working.

Participants were taught how to use thermometers so that accurate temperatures were measured during household visits. Participants were given a chance to practice using thermometers and were instructed to conduct household visits over a period of 4 - 5 days. Youth were present to assist the community health workers during training sessions. In addition, youth were present during household visits to assist with digital data collection.

To reduce any risks of infection, instructions were provided that household visits were to be conducted outside the houses. In addition, hand washing was to occur at the start and at the end of the visit, and there was universal mask wearing by all present. Data collection was done digitally by CHWs and youth.

In parallel, we trained CHAs on how to conduct a community health unit assessment to determine Community Health Unit (CHU) functionality. Three dimensions were assessed: the presence of an active CHW workforce; the presence of active governance structures; and the presence of community health information highlighting the number of households visited, antenatal clinic attendance (ANC), facility based deliveries, and immunization.

Internet data bundles were provided for data collection.

Following the CHU assessment, we ranked CHUs according to the number of CHWs that were active and according to the number of households visited in the previous month.

### Health system level

We conducted training sessions to introduce the digital tool to hospital based health records officers. During the training session, each question was reviewed by facilitators. Furthermore, we emphasized the importance of determining the source of data (primary sources were preferred). Participants were given the chance to ask clarifying questions. Furthermore, participants were given the opportunity to practice using the digital tool.

Targets and deadlines were provided. Internet data bundles were provided for digitized data collection. The health facility digital tool assessed the number and composition of the workforce, the infrastructure available (electricity and running water), availability of personal protective equipment, presence of clinical guidelines, availability of essential maternal and child health services, training in COVID19 diagnosis and management, and availability of essential equipment (pulse oximeters, COVID19 testing kits, and xray), supplies (oxygen, vaccinations), and essential medicines (antibiotics, intravenous fluids etc…).

### Statistical analyses

We calculated frequencies for community level and health system level variables. Data were entered into excel data sheets, and SPSS 20.0 was used to analyze the data.

### Ethical review approvals

received from the University of Nairobi Ethics Review Committee and Jaramogi Oginga Odinga Teaching and Referral Hospital Ethics Review Committee.

### Patient and public involvement

Patients and/or the public were not involved in the design, or conduct, or reporting, or dissemination plans of this research.

## Results

### (1) Community level

We visited a total of 19’474 households.

Table 1 summarizes the prevalence of cough and objectively measured temperatures at household level in 5 subcounties in Siaya. We found that the prevalence of cough ranged from 0.2 to 0.8%; and the prevalence of fever ranged from 1.4% to 4.3%. Forty one households with objective fever were followed clinically. None of these households progressed to COVID-19; and none met case definition.

**Table 1:** Baseline prevalence of cough and summary of objectively measured temperatures at household level in 5 subcounties:

|  |  | ALEGO | UGUNJA | GEM | RARIEDA | UGENYA |
| --- | --- | --- | --- | --- | --- | --- |

|  |  | CHU<br>active | CHU<br>Inactiv<br>e | CHU<br>activ<br>e | CHU<br>Inactiv<br>e | CHU<br>activ<br>e | CHU<br>Inactiv<br>e | CHU<br>activ<br>e | CHU<br>Inactiv<br>e | CHU<br>activ<br>e | CHU<br>Inactiv<br>e |
| --- | --- | --- | --- | --- | --- | --- | --- | --- | --- | --- | --- |
|  |  | n(%) |  | n(%) |  | n(%) |  | n(%) |  | n(%) |  |
| Baseline<br>complaints<br>of cough | Cough | 25<br>(0.2) | 13<br>(0.6) | 2<br>(0.2) | 1<br>(0.6) | 19<br>(1.0) | 1<br>(0.4) | 6<br>(0.6) | 14<br>(0.8) | 61<br>(0.5) | 6<br>(0.5) |
|  | *Chi-square (p*<br>*value)* | **141.307 (0.000)**<br>**\*\*** |  | **25.394 (0.000)**<br>**\*\*** |  | **218.508**<br>**(0.000) \*\*** |  | **24.032 (0.001)**<br>**\*\*** |  | **227.665**<br>**(0.000) \*\*** |  |
| Temperatu<br>re | Normal | 18331<br>(98.6) | 4573<br>(97.3) | 1710<br>(95.7<br>) | 720<br>(98.1) | 3096<br>(98.5<br>) | 548<br>(98.0) | 1049<br>(97.6<br>) | 1555<br>(97.8) | 1532<br>1<br>(98.4<br>) | 2295<br>(98.0) |
|  | High | 264<br>(1.4) | 129<br>(2.7) | 76<br>(4.3) | 14<br>(1.9) | 48<br>(1.5) | 11<br>(2.0) | 26<br>(2.4) | 35<br>(2.2) | 256<br>(1.6) | 46<br>(2.0) |
|  | *Chi square*<br>*significant*<br>*value* | **0.000\*\*** |  | **0.004\*\*** |  | 0.443 |  | 0.713 |  | 0.260 |  |

CHU- Community Health Unit, \*\* $p < 0.01$ , \* $p < 0.05$

Table 2 summarizes baseline household knowledge of COVID-19 preventive measures and adherence to these preventive measures. We found that community knowledge of preventive measures and adherence to preventive measures were high.

**Table 2:** Baseline household knowledge of COVID-19 preventive measures and adherence to preventive measures:

| Variable | % |
| --- | --- |
| Knowledge about frequent handwashing | 93% |
| Knowledge about need for physical distancing | 67% |
| Knowledge about need for wearing a mask | 90% |
| Do not know any COVID-19 preventing measures | <1% |
| Had not received a guest from outside Siaya | 97.8% |
| Had not visited a county outside Siaya | 98.1% |
| Had washed their hands > 5 times | 94% |
| Had not gone out in public in the last week | 53% |
| Had used a face mask when out in public | 74% |

Table 3 summarizes baseline reported use of maternal health services. We found that antenatal clinic attendance and delivery in health facilities were high.

**Table 3:** Use of maternal health services:

| Service | % |
| --- | --- |
| Antenatal clinic attendance during last pregnancy | 78.6% |
| Delivery at health facility during last pregnancy | 85% |

#### Baseline COVID-19 training and use of pulse oximetry and thermometers

At baseline, CHWs did not have access to thermometers nor pulse oximeters. They had not received training in COVID19 case management. Furthermore, before the intervention CHWs had not carried out door-to-door screening for COVID-19 symptoms, contact tracing, and had not assessed community member knowledge of COVID19 preventive measures. Furthermore, use of digital tools and contactless thermometers were new skills for CHWs and youth.

### (2) Health facility level

An assessment of 152 facilities revealed that 93.6% of health facilities in Siaya were lower level facilities (levels 1 and 2 i.e. dispensaries and health centers); critical care capacity was limited. Siaya county referral hospital had 9 ICU beds with 2 ventilators; and the majority of facilities were staffed by nurses.

Table 4 summarizes the workforce composition in Siaya facilities. We found that the majority of facilities are staffed by nurses.

**Table 4:** Workforce composition in Siaya facilities:

| Cadre | % of facilities with cadre |
| --- | --- |
| General medical doctors | 20.3% |
| Nurses | 90.4% |
| Clinical officers | 65.8% |

Table 5 summarizes basic critical care capacity in facilities and the availability of essential maternal and child health services. We found that the majority of facilities did not have oxygen capacity and pulse oximeters. In addition, we found that the majority of facilities had antenatal and immunization services.

**Table 5:** Basic critical care capacity and availability of essential maternal and child health services.

| Variable | % |
| --- | --- |
| Oxygen available | 40.5% |
| Pulse oximeter available | 13.5% |
| Antenatal care | 93.6% |
| Immunization | 90.7% |

Three public facilities in Siaya had functional operating rooms (1.9%).

In terms of basic infrastructure and equipment necessary for infection prevention and control: 94% of facilities had clean running water; 98.2% had soap for handwashing; 94% had disposable gloves; and 76.9% had masks. However, testing capacity for COVID-19 was very limited: more than 94% of facilities did not have COVID-19 testing kits; and 93.6% did not have functional x-ray machines.

Furthermore, none of the health care workers had received training in basic critical care, and none of the health care workers had received training in COVID19 case management. A charge nurse in Rarieda recently stated: “if a patient with COVID19 came today, we would not know what to do.”

Essential maternal and child health services remained available: 93.6% of facilities continued to provide antenatal care, and >90.7% continued to provide immunization services.

## Discussion

We completed a baseline assessment in 5 Siaya subcounties, reaching 20’000 households and a population of 100’000. At the health system level, we evaluated 152 health facilities to evaluate the health workforce, essential infrastructure and equipment needed for COVID-19 case management, and the availability of essential maternal and child health services.

### The main findings were

We found that the prevalence of cough ranged from 0.2 to 0.8%; and the prevalence of fever ranged from 1.4% to 4.3%; 97% of households had not received a guest from outside Siaya and 98% had not traveled outside Siaya. Knowledge of COVID19 preventive measures was high: 93% of respondents knew about frequent handwashing with soap and water, 67% knew about physical distancing, and 80% knew about mask wearing; <1% reported not knowing any preventive measures. Action to prevent COVID19 was common: 74% used a face mask when going out in public; and 94% of respondents washed their hands more than 5 times daily.

Health seeking behavior was maintained: 78.6% of women attended antenatal clinics, and 85% delivered at health facilities during their last pregnancies. Households with fever did not develop COVID-19 symptoms and never met the case definition for COVID-19.

At baseline, because of national lockdown measures, there was limited travel in and out of Siaya county. Siaya residents showed a high level of knowledge of COVID-19 prevention measures; and they showed high adherence to instructions to wear masks, and wash their hands frequently. These findings demonstrate effective communication by the ministry of health mostly through CHWs, radio and telephone messages that reached households during the early phase of the COVID-19 pandemic. The low prevalence of fever, and the non progression of households with fever shows that at baseline, community transmission was likely low due to travel restrictions. Establishing the baseline prevalence of fever and respiratory symptoms is useful for future comparisons to determine changes from baseline, and to determine need for more intense policy action to prevent and contain COVID-19 cases.

A review of the literature does not show other baseline measurements of the prevalence of fever and respiratory symptoms in Siaya Kenya during COVID-19. However, seroprevalence for COVID-19 antibodies was measured; positivity was found to be 5.6% (neighboring Kisumu had a positive seroprevalence rate of 5.5% between April and June 2020) (15).

Although Odhiambo et al report a 50.7% reduction in use of HIV services in 60 MOH clinics in Kenya since March 2020, reported use of maternal health services remained high during our baseline assessment suggesting effective promotion of maternal health services by the MOH, and health care workers at community and health system levels (16). Maintained use of essential maternal health services is a significant gain that should be maintained throughout the COVID-19 response. Other pandemics including the Ebola outbreaks in West and central Africa resulted in the erosion of access to essential health services (17-24). A multicountry assessment of the impact of COVID-19 on access to health services demonstrated a reduction in the ability of slum residents to seek health care for non COVID-19 conditions in Bangladesh, Kenya, Nigeria, and Pakistan (25).

At baseline, we found that CHWs and youth had not received prior training in COVID-19. In addition, they had not received prior training in use of digital tools to complete data collection for COVID-19 screening, contact tracing, and case management. Prior evidence has demonstrated the reliability and validity of community based data collected by CHWs (10, 11). In this study, we demonstrated the feasibility of using digitized household surveys for more accelerated decision making by policy makers. In this intervention, CHWs were instrumental in baseline data collection and promoting health behavior that prevented and contained COVID-19 cases, and promoted maintained use of essential health services.

At the health facility level, an assessment of 152 facilities revealed that critical care capacity was limited in terms of the workforce, infrastructure, equipment, and essential supplies such as oxygen. In addition, the diagnostic capacity for COVID-19, and clinician knowledge of COVID-19 case management was limited. The majority of health facilities were lower level facilities and were staffed by nurses. Two ventilators were available for the population of 993’000. A recent assessment of the national ICU capacity in Kenya also demonstrated limited capacity; they found that only 58% of hospital beds in Kenya had an oxygen supply and that only 22 out of the 47 counties had at least 1 ICU unit (26). Essential maternal and child health services remained available.

As a result of the above findings, any intervention addressing gaps in COVID-19 preparedness and response should prioritize the capacity building of the nursing and clinical officer workforce; the training should include improving knowledge and skills in COVID-19 case management and basic critical care; and the provision of pulse oximeters as a critical clinical monitoring tool should be prioritized. 1.97% of hospitals in Siaya had operating room theaters; therefore, repurposing operating rooms to build critical care capacity is not a viable solution for the vast majority of hospitals in Siaya; and may even jeopardize access to life saving surgical care.

In terms of basic infrastructure and equipment, 97% of facilities had clean running water; and 98% had soap for handwashing. However, diagnostic capacity was low: 94.2% of facilities did not have COVID-19 testing kits, and 93.6% did not have functional x ray machines. Consequently, future interventions should focus on emphasizing handwashing as part of IPC and emphasize using clinical signs and symptoms and pulse oximetry to diagnose and subsequently manage COVID-19 in health facilities.

At the health system level, safety of healthcare workers has emerged as a priority issue (27). Our baseline assessment showed that early in the pandemic, Siaya had basic PPE: 94% had disposable gloves, and 76.9% of facilities had masks. The challenge will be maintaining these commodities as demand increases during the course of the pandemic.

### This study has several strong points

It is the first integrated and comprehensive assessment of pandemic preparedness in Siaya - a rural county in Kenya. It was performed early during the pandemic allowing policy makers to design interventions that were context specific and addressed the gaps that came to light during the baseline assessment. It demonstrated the feasibility of using digital tools for data collection and analysis during a pandemic. This baseline assessment provides evidence that will serve as a starting point for future comparisons. Our sampling frame was broad at community and health system levels; therefore, significant selection bias was unlikely.

We developed and implemented an approach that worked in the rural context. Forty four percent of the global population remains rural, and many governments and ministries of health will need to assess preparedness and plan interventions to adequately respond to COVID-19 in their rural populations. This study provides an example of how this can successfully be done with CHWs, youth, and digital tools (4). This is particularly important for African countries such as Kenya where rural populations make up up to 72% of the population and there is an established community health strategy (4). Moreover, in the current context of multiple and persistent waves of COVID-19 infections, such an exercise by ministries of health is urgent given the limited resources available in rural health systems and their vulnerability to being rapidly overwhelmed.

### This study has several limitations

1. We had delays in data acquisition using digital tools because of poor network coverage in rural Siaya. Some CHWs needed additional technical support when using digital tools. This may have resulted in missed data points from household visits. To minimize this, we instructed all those involved in data collection to have back up paper versions of tools to complete and transfer to the digital system when the network coverage improved.
2. These baseline findings are likely to change over time with changes in travel restrictions, and increased demands for commodities related to COVID-19 management (in particular PPE and oxygen). Potential healthcare worker strikes will also affect the workforce composition and density.
3. Some of our data were retrospective in nature and therefore vulnerable to multiple sources of bias including: recall bias and misclassification.

## Conclusion

At baseline, the prevalence of fever and respiratory symptoms was low suggesting we had not entered the active community transmission phase in June of 2020. Our assessment revealed strengths and limitations at both community and health systems levels. Community knowledge about COVID-19 prevention was high and adherence to preventive measures was high; however, more training was needed to strengthen CHW knowledge and skills to optimize COVID-19 case management.

At the health system level, we documented the availability of maternal and child health services, and basic commodities and infrastructure for IPC were present; however, critical care capacity, COVID-19 diagnostic capacity, and knowledge of COVID-19 cases management were limited.

Future interventions should maintain and build on the strengths highlighted and strengthen areas of weakness in particular, CHW and clinician knowledge and skills in COVID-19 case management, basic critical care capacity, and coverage of basic equipment such as pulse oximeters.

Lastly, this study provides an example of how to successfully carry out an integrated rural health system baseline assessment of pandemic preparedness using CHWs, youth, and digital tools. Such an approach would be useful for any country experiencing COVID-19 with a significant rural population and an established community health strategy.

## Data Availability

There are no contractual agreements limiting access to datasets; datasets will be made available upon submission of an approved proposal.

## a. Contributorship statement

NK wrote the 1st draft. All co-authors provided input on subsequent drafts. All authors read and approved the final manuscript.

NK: Conceptualization, Funding Acquisition, Investigation, Methodology, Project Administration, Writing – Original Draft Preparation.

DK: Conceptualization, Methodology, Writing – Review & Editing

KO: Author contribution(s): Project Administration, Writing – Review & Editing

PO: Conceptualization, Data acquisition, Writing – Review & Editing

MK: Conceptualization, Data acquisition, Writing – Review & Editing

SA: Data acquisition, Writing – Review & Editing

MT: Conceptualization, Methodology, Writing – Review & Editing

AH: Conceptualization, Methodology, Writing – Review & Editing

## c. Data sharing statement

There are no contractual agreements limiting access to datasets; datasets will be made available upon submission of an approved proposal. Full protocols will be available upon request.

